# Successful Tracheal Extubation within Eleven Minutes in the Post Anesthesia Care Unit in Children after Adenotonsillectomy

**DOI:** 10.1101/2020.09.04.20188086

**Authors:** Deepak Gupta, Matthew Ryan Tukel, Divya Mukhija, Edward Kaminski, Maria Markakis Zestos

## Abstract

**Background:** Some pediatric centers prefer to extubate their patients in the operating rooms (ORs) while others prefer post-anesthesia care units (PACUs) for the same.

**Objectives:** To share our retrospective experience of 214 pediatric adenotonsillectomy (T&A) patients’ cohort extubated in our pediatric PACU during a seven-month retrospective study-period.

**Materials and Methods:** After institutional review board approval for retrospective chart review, institutional electronic surgical database was used to identify patients who underwent T&A and the peri-anesthetic records were obtained from patients’ electronic medical records and/or from hospital paper records.

**Results:** Patients’ tracheas were extubated in average 11 minutes (standard deviation 8 minutes) after arrival to PACU care and only one patient required tracheal re-intubation. Patients were ready for discharge from PACU in average 56 minutes (standard deviation 20 minutes) thus averaging only 44 minutes (standard deviation 20 minutes) after their tracheas had been extubated.

**Conclusion:** Summarily for re-validating or refuting our results, institutions can prospectively create PACU extubation quality improvement projects to discern if tracheal extubation in PACU of all or some pediatric surgical patients is beneficial when their rapid turnover surgeries warrant anesthesia providers to not attempt their tracheal extubation in ORs.

## Introduction

Ideal anesthesia work sites for pediatric post-anesthesia endotracheal extubation have recently been in debate [1–10]. Some pediatric centers prefer to extubate their patients in the operating rooms (ORs) while others prefer post-anesthesia care units (PACUs) for the same. In our view, there are primarily three considerations namely patient safety, work flow logistics and hospital-costs/patient charges. Essentially, the location for extubation depends on PACU availability, patient condition, and the need for rapid turnover in operating rooms. For these reasons, at our children’s hospital, if space is available in the PACU, we often extubate in PACU. Tracheal extubation can be performed by appropriately trained PACU nurses or by the anesthesia team who can remain with the patient in the PACU until extubation. However, if the PACU is full or the patient condition necessitates it, we often extubate in the OR. Thus, our initial hypothesis was that there would be no difference in patient outcomes whether pediatric patient’s tracheas were to be extubated in the OR vs. in the PACU, but that there would be potential cost savings with extubation in the PACU, from improved operating room throughput.

## Materials and Methods

Institutional review board approval for retrospective chart review was taken wherein the retrospective chart review was designed to ascertain the differences in time to patient recovery/discharge, the incidence of post-anesthesia patient events based on location of pediatric extubation (in OR vs. in PACU), and cost differences that arise given the location of extubation and time of recovery. This review analyzed pediatric patients under the age of 18 who had come for elective adenotonsillectomy (T&A) from July 2009 to June 2015 at our hospital. However, due to logistic reasons and research personnel’s time-limitations, we were able to extract, analyze and tabulate data only for a period of seven months in 2014-2015 study-period wherein almost none of the T&A patients were extubated in ORs. Thus, we are just sharing our experience of 214 pediatric T&A patients’ cohort extubated in our pediatric PACU during this seven-month retrospective study-period.

Institutional electronic surgical database was used to identify patients who underwent T&A and the peri-anesthetic records were obtained from patients’ electronic medical records (EMR) and/or from hospital paper records. Once the appropriate records had been obtained, we extracted the patient’s age, sex, height, weight, anesthesia start and stop time, duration of surgery (incision to closure), duration of PACU stay (PACU entry to PACU exit/readiness), exact time of extubation if the patient’s trachea was extubated in the OR, and exact time of extubation if patient’s trachea was extubated in the PACU. Finally, we obtained data regarding the occurrence of patient events during the procedure including laryngospasm, bronchospasm, re-intubation, non-invasive positive pressure ventilation, post operative ventilation, history of asthma or reactive airway disease, recent upper respiratory tract infections (< 6 weeks), and any other significant procedural events.

## Results and Discussion

The average age of these 214 pediatric T&A patients was 7 years 3 months (standard deviation 3 years 10 months). Fifty one percent were females and 49% were males. The average height was 124 centimeters (standard deviation 22 centimeters) and average weight was 31 kilograms (standard deviation 19 kilograms). While 43% had preoperative history of asthma/reactive airway disease, only 8% had preoperative history of recent upper respiratory tract infections (< 6 weeks).

Anesthesia duration averaged 56 minutes (standard deviation 13 minutes) and surgery duration averaged 32 minutes (standard deviation 11 minutes).

Patients’ tracheas were extubated in average 11 minutes (standard deviation 8 minutes) after arrival to PACU care. Although patients were ready for discharge from PACU in average 56 minutes (standard deviation 20 minutes) thus averaging only 44 minutes (standard deviation 20 minutes) after their tracheas had been extubated, they actually got discharged from PACU in average 3.5 hours (standard deviation 17.3 hours). This is in part due to a hospital policy to keep T&A patients for a minimum of 1.5 hours postoperatively and until they can drink adequately to stay hydrated.

During PACU stay, 9% of patients received bronchodilator therapy (albuterol, ipratropium bromide, or racemic epinephrine) even though none of the patient charts had documented laryngospasm or bronchospasm. Two patients did have documented stridor and one patient required tracheal re-intubation but none of the patient charts had documented use of non-invasive positive pressure ventilation or intensive care unit admission for postoperative mechanical ventilation.

Although hospital costs and patient charges were not calculated, it can be easily said that an average 11-minute OR time was saved by allowing patients’ tracheas to be extubated in PACU and correspondingly per-patient healthcare cost savings would accrue from spending less time in the higher-cost/charge OR and more time in the lower-cost/charge PACU.

## Conclusion

In summary, institutions can prospectively create PACU extubation quality improvement projects wherein they can re-validate or refute whether tracheal extubation in PACU of all or some pediatric surgical patients is beneficial when their rapid turnover surgeries warrant anesthesia providers to not attempt their tracheal extubation in ORs.

## Data Availability

Data as related to results is available.

## Acknowledgements

The authors are thankful to Dr. David Wolma for his contributions to this project at the time of his pediatric anesthesiology fellowship at Children’s Hospital of Michigan, Detroit Medical Center, Detroit, Michigan, United States.

## Notes

**Conflicts of Interests**: None

### Competing Interest Statement

The authors have declared no competing interest.

### Funding Statement

Nothing to report.

### Author Declarations

Institutional Review Board Wayne State University Detroit MI

